# Personal Resilience, Social Support, and Organizational Support Impact Burnout among Nurses During COVID-19

**DOI:** 10.1101/2022.01.05.22268637

**Authors:** Hanan Daghash

## Abstract

**Background:** Nurses have been under heavy workloads since the outbreak of COVID-19 and are at a high risk of infection, leading to a high level of psychosocial risk. This can adversely affect nurses both psychologically and physically. Burnout is caused by prolonged stress during work. In the nursing profession, burnout is common, potentially affecting the well-being of nurses and their productivity. The identification of factors that may contribute to maintaining mental health and reducing burnout among frontline nurses during a pandemic is essential.

**Purpose:** The purpose of this study was to explore how personal resilience, social support, and organizational support impact burnout among frontline staff nurses.

**Methods:** This study involved 129 registered nurses from a COVID-19 designated hospital using four standardized scales.

**Results:** The mean age of the respondents was 29.46 years (standard deviation = 4.89). The mean number of years respondents worked in this organization was 5.60 years and the nursing profession was 4.16 years. Most of the respondents were female and held a bachelor’s degree in nursing. Multiple regression analysis was performed to predict burnout. Burnout was statistically significantly predicted by the multiple regression model (*R*^*2*^ = .420, *F* (3, 125) = 10.941, *p* < .0001; adjusted *R*^*2*^ = .406). Personal resilience, social support, and organizational support added statistically significantly to the prediction of burnout (*p* < .05).

**Conclusion:** Findings from multiple regression analysis showed that nurses with low resilience and those who perceived inadequate social and organizational support had a higher risk of reporting more burnout. As a result of a bivariate analysis, there was no significant correlation between nurse variables and burnout level, except for age, which was negatively correlated with burnout level. Accordingly, young nurses tend to experience burnout, and nurse directors and managers must address this problem.

## Introduction

In December 2019, a coronavirus related to severe acute respiratory syndrome (SARS-CoV-2) and a related disease (COVID -19) was identified in Wuhan, China [1]. The SARS-CoV-2 infection has gone significantly faster than other recent pandemic diseases (SARS, MERS) [2]. Because the seroprevalence of SARS-CoV-2 antibodies among healthcare workers is stronger than that of the general population globally, they are a particularly high-risk group for infection with SARS-CoV-2 [3]. According to a meta-analysis that included data until May 8, 2020, nurses (259 of 1024) contributed 25.3 percent of COVID-19 mortality among healthcare professionals, with nurses having the highest chance of disease [4].

Nurses who provide care to patients infected with SARS-CoV-2 are at risk of developing psychological problems, as well as other mental health problems [5]. Healthcare workers may suffer from mental distress due to a variety of aspects, including personal protection equipment, an increasing number of confirmed and suspected cases, a lack of specific medications, burdensome workloads, extensive media coverage, and a sense of not being sufficiently supported. Research published in 2020 revealed a striking prevalence of post-traumatic stress disorder (PTSD) among healthcare workers after disasters [5]. Researchers cautioned, however, that studies focusing on psychological disorders, such as burnout, are lacking. Nurse burnout is a major problem during the current COVID-19 epidemic, according to a newly published systematic review; consequently, nurses need better training to handle stressful situations during the pandemic [6]. In the near future, identifying risk factors for burnout could be an important tool in helping nurses and health systems better respond to subsequent waves of COVID-19.

Burnout has become increasingly recognized as a major occupational problem that impedes the physical and psychological well-being of healthcare providers worldwide. Burnout is a condition of mental emotional, fatigue, and physical problems caused by long-term experience in emotionally challenging work situations [7]. In 2019, according to the World Health Organization, burnout is characterized by “feelings of energy depletion or exhaustion; increased mental distance from one’s work or feelings of negativism or cynicism about one’s work; and reduced professional effectiveness.” In part, high levels of burnout may continue during the coronavirus pandemic (COVID-19), when healthcare providers are overwhelmed with heavy workloads in delivering healthcare services and nurses are at risk of infection [4]. It has been shown that burnout can lead to suboptimal patient care in addition to being personally harmful [9].

The risk of burnout can be reduced if nurses are aware of the risks and prepared for possible occupational stress. In addition to helping nurses cope with and adapt to difficult occupational conditions, resilience may prevent mental disorders such as burnout [10]. A study of the Ebola response revealed that situational understanding and other preventative measures enhanced mental resilience among healthcare workers [11]. It is thought that social support provided by colleagues, supervisors, friends, and families is crucial in helping nurses cope with various stressors in the occupational environment. Multiple studies have noted the positive effects of social support on burnout [12], [13] t. Providing adequate social support was considered important in helping nurses manage stressful situations, such as COVID-19 [6]. Organizational support, or the extent to which an organization commits to providing resources, reinforcing, encouraging, and communicating with its members, is a critical determinant of organizational success [14]. Higher levels of organizational support may help to reduce the impact of multiple workplace stressors and protect employees from stress caused by disasters, catastrophes, and new diseases [15]. Burnout feelings of being emotionally drained and lacking emotional resources. The significance of personal resilience, social support, and organizational support in preserving the health and well-being of nurses in stressful situations was investigated in this study. However, the evidence for this relationship cannot be conclusively shown. This study aimed to determine the role of personal resilience, social support, and organizational support in predicting burnout among nurses assigned to a COVID-19 designated hospital.

## Methods

### Research Design

A cross-sectional design was used in this study. In this study, nurses were asked to fill out a voluntary questionnaire from every department and ward in the hospital. Data were collected using an online self-report survey via Google Survey between February 13, 2021, and March 5, 2021.

### Participants and Study Setting

A convenience sample of 129 nurses from a public hospital in Tabuk, Saudi Arabia, was enrolled in this study. The hospital was designated to treat patients who were positive for SARS-CoV-2 antibodies. A questionnaire was sent to the participants electronically. The size of the sample was initially determined using the power estimates for three predictors in multiple regression to achieve 80 percent power with an alpha of 0.05, and a minor effect size of 0.05. Only 129 nurses responded to the invitation to participate in the study (58%) out of a total of 220 participants.

### Research Instrument

This study used a self-administered questionnaire containing two sections. A total of seven demographic questions were included in Section I, including age, gender, level of education, wards, and the number of years of work experience.

The Copenhagen Burnout Inventory (CBI), the Perceived Organizational Support Questionnaire (POS), the Perceived Social Support Questionnaire (PSSQ) and the Brief Resilient Coping Scale (BRCS) are included in Section II.

The Copenhagen Burnout Inventory was used to assess the degree of burnout Inventory (CBI) was utilized. The CBI consists of 19 items rated on a 5-point Likert scale. Five items were included in the first domain, which addressed personal burnout. Six items were included in the second domain, which was based on work-related burnout. In the third domain, there were eight items based on client-related burnout. There are 19 items on the CBI and a 5-point Likert scale: 100 (always), 75 (often), 50 (sometimes), 25 (rarely), and 0 (never/almost never). Low or no risk of burnout was indicated by a score below 50, moderate or high risk of burnout was indicated by a score between 50 and 74, and a high risk of burnout is indicated by a score of more than 75 [16].

The Brief Resilient Coping Scale (BRCS)15 is used to assess nurses’ ability to cope after stressful events. On a scale of 0 to 5, nurses were asked to rate each of the four items (0 (does not describe me at all) to 5 (describes me very well). There were three groups of mean scale scores [17]: low resilience (1.00-2.99), medium resilience (3.00-4.30), and high resilience (4.31-5.00).

Nursing perceptions of the level of social support that they receive from other individuals in stressful situations were assessed using a PSSQ designed by [18]. On a 5-point Likert scale, a nurse answered the PSSQ by expressing their level of agreement with each of the six items (1 = strongly disagree to 5 = strongly agree). Low social support (1.00-2.99), moderate social support (3.00–4.30), and strong social support (4.31–5.00) were the three scale categories.

According to Eisenberger et al.[19], the well-being scale established by them was used to evaluate nurses’ views of workplace values and organizational support. In general, the mean scores on the scale can be categorized into three groups: low organizational support (1.00–2.99), moderate organizational support (3.00–4.30), and high organizational support (4.31–5.00) [20]. According to its positive associations with work satisfaction, quality of life, and psychological well-being, the measure had excellent criterion validity [20].

The questionnaire was designed to be publicly available in English and translated into Arabic. In this study, Cronbach’s α coefficients were 0.837, 0.817, 0.871, and 0.944 for personal resilience, social support for organizational support, and burnout, respectively. In terms of internal consistency, the questionnaires were considered reliable because all α -coefficients obtained exceeded 0.70.

### Ethical considerations

This study was approved by the Institutional Review Board (IRB) of the Tabuk region’s General Director of Health Affairs (reference number: TU-077/020/064). Participants were sent a copy of the questionnaire and an information sheet explaining the purpose and aim of the study, the voluntary nature of participation, and the confidentiality of respondents’ responses.

### Data analysis

The data were quantified using frequencies, standard deviations (SDs), and means. The correlations between demographic variables and key variables of the study were tested using Student’s t-tests, Pearson’s correlation coefficients, and analysis of variance. A regression model (enter method) was developed based on variables that were significantly correlated with the dependent variable. For statistical tests, a p value of 0.05, was considered statistically significant. The data for this study were analyzed using the SPSS statistical software version 23 for Windows 22.

## Results

A total of 129 nurses completed the questionnaire. The sample age was 29.46 years (SD = 4,89), whereas mean years spent in both the current organization and nursing profession were 5.60 years and 4.16 years, accordingly. More than 80 percent of the respondents were female (83.7%), and eighty-three percent had a bachelor’s degree in nursing (83.7%). Table 1 presents the nurses’ specific characteristics.

**Table 1.**
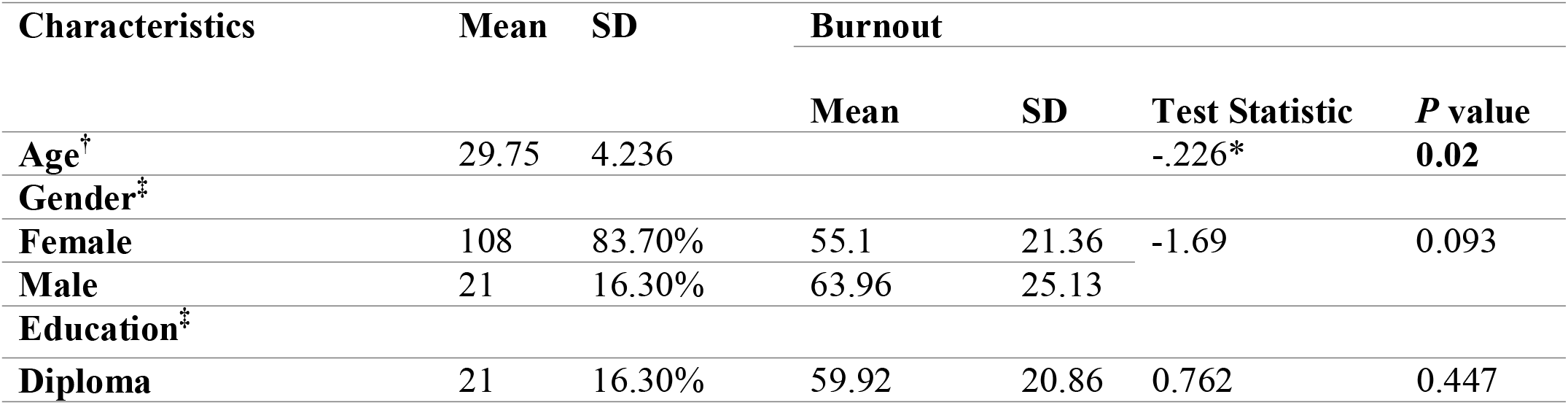
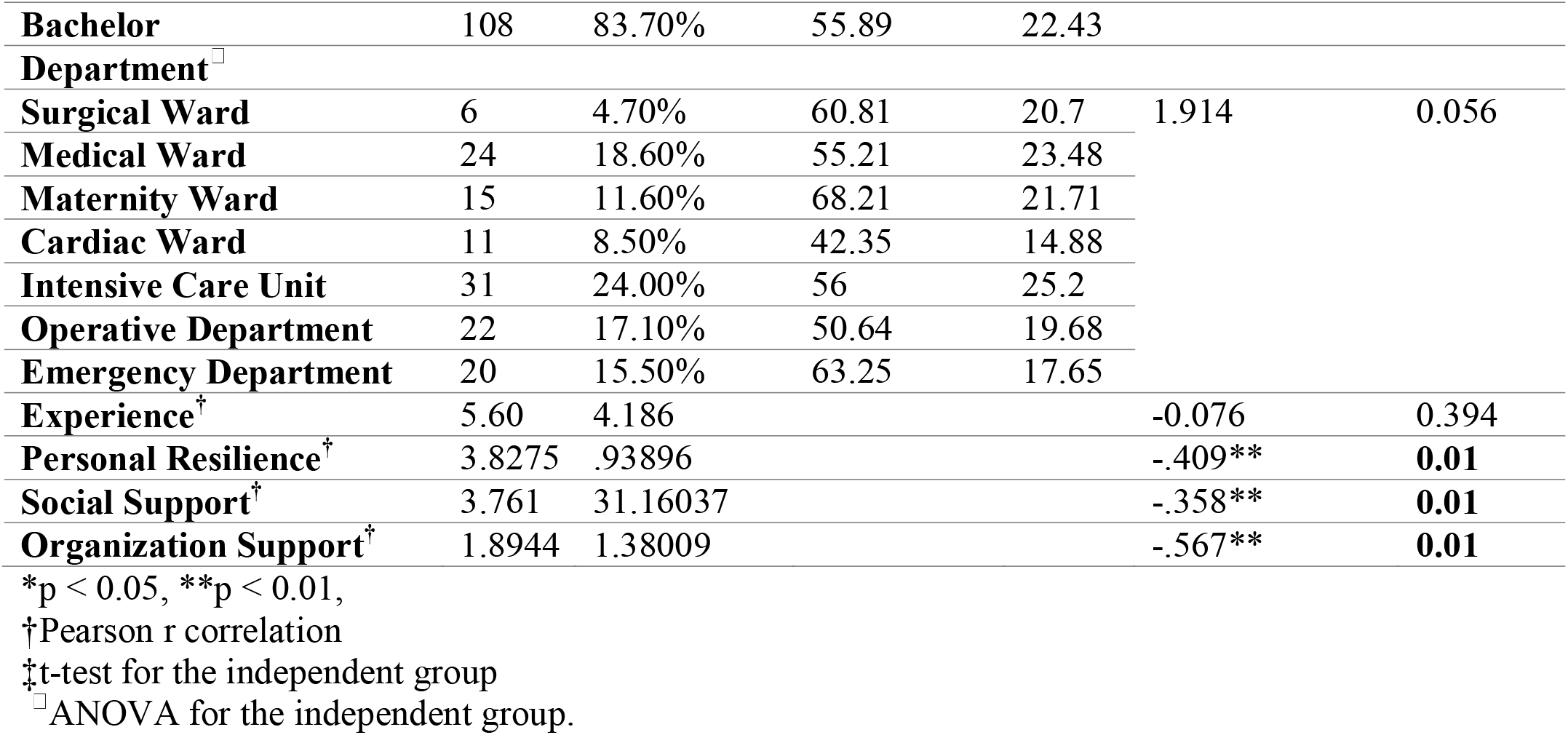
Nurses’ demographic and professional information and its relationship with burnout mean score.

According to a bivariate analysis, there was no significant correlation between nurse variables and the composite score of burnout scale, except for age, which was negatively related to burnout level. The Pearson correlation analysis identified strong negative relationships between burnout and resilience (*r* = -0.409, *p* < 0.01), social support (*r* = -0.358, *p* < 0.01), and organizational support (*r* = -0.567, *p* < 0.01; Table 1). Thus, this would imply a decrease in the organization’s social support and a decrease in resilience, thereby increasing burnout levels.

Scale scores for the Brief Resilient Coping Scale were moderate (3.83, SD = 0.938), social support perceptions were rated moderate, whereas organizational support was rated poor and below average, according to rating scales of 3.67 and 1.803, respectively.

Personal resilience, social support, and organizational support were utilized in this research to predict burnout using multiple regression analysis. Partial regression plots and a plot of studentized residuals against the expected values revealed linearity. Burnout was statistically predicted by the multiple regression model, with *R*^*2*^ = .420, *F* (3, 125) = 10.941, *p* < .0001; adjusted *R*^*2*^ = .406. The models indicated significant negative associations between social support, organizational support, and personal resilience and burnout. The prediction was statistically significant for all four factors (*p* < 0.05). Table 2 presents the regression coefficients and standard errors (below). In other words, lower personal resilience, social support, and organizational support values were linked to greater burnout.

**Table 2.**
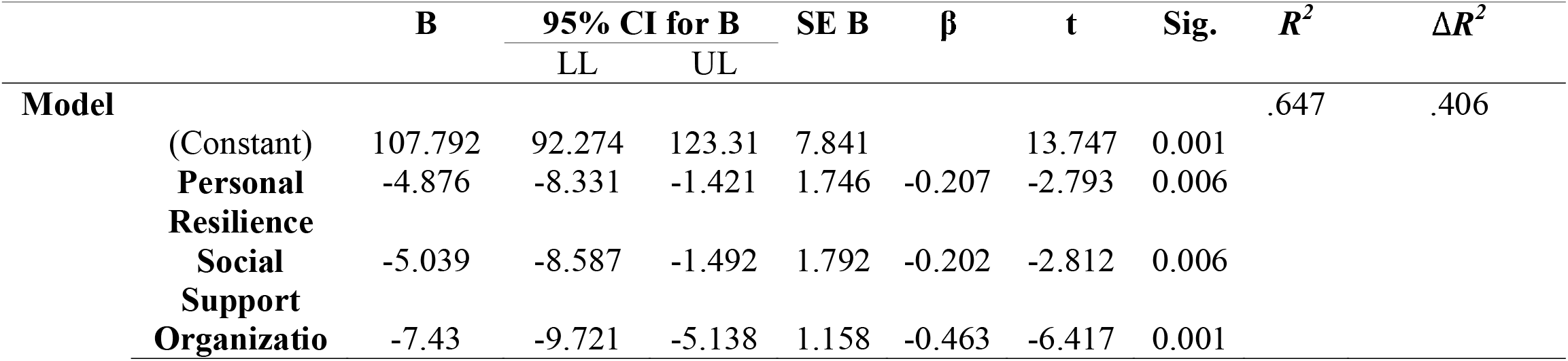
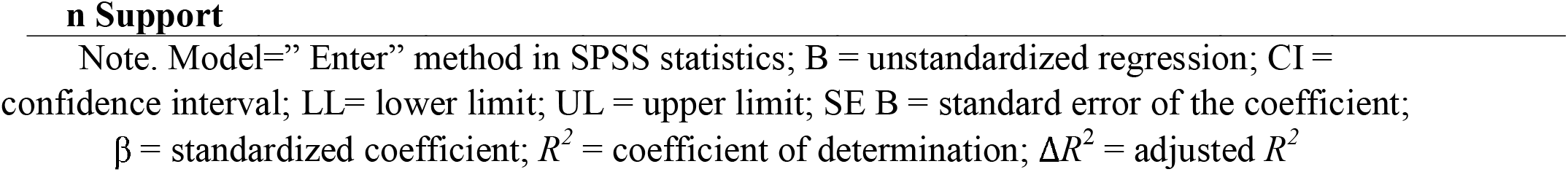
Influence of personal resilience, social support, and organizational support on burnout among nurses.

|  | **B** | **95% CI for B** |  | **SE B** | **$\beta$** | **t** | **Sig.** | **$R^2$** | **$\Delta R^2$** |
| --- | --- | --- | --- | --- | --- | --- | --- | --- | --- |
|  |  | LL | UL |  |  |  |  |  |  |
| **Model** |  |  |  |  |  |  |  | .647 | .406 |
| (Constant) | 107.792 | 92.274 | 123.31 | 7.841 |  | 13.747 | 0.001 |  |  |
| **Personal Resilience** | -4.876 | -8.331 | -1.421 | 1.746 | -0.207 | -2.793 | 0.006 |  |  |
| **Social Support** | -5.039 | -8.587 | -1.492 | 1.792 | -0.202 | -2.812 | 0.006 |  |  |
| **Organizational Support** | -7.43 | -9.721 | -5.138 | 1.158 | -0.463 | -6.417 | 0.001 |  |  |

## Discussion

As a result of COVID-19’s higher frequency probability and global dissemination, healthcare practitioners are put under a great deal of burden. Nurses caring for infected patients are exposed to a constant risk of infection. They are afraid of their personal health and the health of their close relatives [5]. Nurses may suffer from serious psychological and mental problems as a consequence of such situations, which may lead to emotional discomfort and burnout, which can then negatively impact their productivity, clinical errors, and behaviors in dealing with patients [9]. In this study, we examined the effects of individual resilience, social support, and organizational support on burnout among frontline nurses working in a high-risk clinical setting as designated by COVID-19. According to this study, during the COVID-19 epidemic, nurses reported moderate degrees of personal resilience and perceived moderate levels of social support, but low levels of organizational support. Additionally, the evidence suggests that nurses with lower levels of personal resilience, social support, and organizational support are more likely to experience burnout.

According to the statistical analysis, there was no significant connection between burnout and demographic factors in this research, such as gender, work department, and years of work experience. This is in line with the results of previous studies [21], [22]. The findings of this study are consistent. However, the present study found that age had a negative relationship with burnout. In accordance with previous research [21], young nurses were found to be associated with burnout due to insufficient support and fewer experiences.

According to this research, lower levels of personal resilience were linked to greater levels of burnout among frontline nurses in a hospital dedicated to COVID-19 patients. The lower burnout rates among nurses who scored higher on the resilience scale suggest that personal resilience plays an important role in protecting individuals against adverse situations [23]. Given this result, it is important to look at methods to improve personal resilience among frontline nurses in order to prevent burnout. According to prior research, greater resilience among nurses corresponds with reduced burnout, psychological distress, depression, anxiety, and compassion fatigue[23], [24]. Additionally, greater resilience has been related to better individual consequences, such as well-being and psychological health. [23], [25].

The present research found that lack of social support was a factor in predicting burnout in the workplace. This study extends the findings of previous studies showing that strong social networks during a pandemic can decrease feelings of isolation in nurses and help them become more resilient[12], [13]. This result may be attributable to the fact that support from families, friends, and colleagues enables nurses to effectively control and avoid negative feelings and emotions, which reduces the risk of burnout.

While the present study is preliminary, the results suggest that nurses working in designated COVID-19 hospitals received very little support or recognition from their organizations during the pandemic. This study also found that a lack of organizational support was predictive of burnout syndrome. These findings are consistent with those of other studies [26], [27]. It is possible that during a pandemic, inadequate support could be provided by the organization, resulting in nurses experiencing frustration and emotional distress, increasing the risk of burnout. We may conclude from these statistics that the organization offers sufficient support for nurses in order for them to maintain excellent conditions for workers, minimize mental and physical resource consumption and burnout at work, and allowing them to struggle and concentrate on their tasks.

Despite the fact that this study provides nursing administrators with evidence to assist nurses in pandemic scenarios, certain limitations were identified. First, the research was limited to a small group of nurses from a particular institution, as the findings cannot be generalized to nurses across the country or the globe. Furthermore, the research design has significant drawbacks, such as the inability to establish causal links between variables. In the future, researchers using both qualitative and quantitative research methods may be able to collect information from participants that cannot be obtained through self-report measures. To determine the effectiveness of resilience management programs and other strategies, rigorous studies using rigorous methods (such as randomized control trials) would need to be conducted. Moreover, during the epidemic, nurses are likely to benefit from mindfulness and/or cognitive behavioral therapy[28]. Ideally, future research should explore individual factors (e.g., self-efficacy, coping skills, and hardiness), supervisors’ factors (e.g., nursing schedules and leadership styles), and organizational factors (e.g., workload, healthcare staffing levels, resource availability, hospital size, and number) play a role in nursing burnout during pandemics.

## Conclusion

A greater degree of burnout was anticipated for nursing workers working in a high-risk, COVID-19-designated hospital with low levels of personal resilience, low levels of social support, and low levels of organizational support. Research conducted during the epidemic supported this result. While the study has several limitations, it concludes that nurses’ ability to maintain psychological and mental health, as well as reducing their risk of burnout syndrome, can be enhanced by maximizing resilience, providing adequate social support, and maintaining good organizational support during the COVID -19 crisis.

## Data Availability

All data produced in the present study are available upon reasonable request to the authors

